# Impact of Anemia on Outcomes and Resource Utilization in Patients with Myocardial Infarction: A National Database Analysis

**DOI:** 10.1101/2023.02.28.23286603

**Authors:** Aravdeep S. Jhand, Waiel Abusnina, Hyo Jung Tak, Arslan Ahmed, Mahmoud Ismayl, S. Elissa Altin, Matthew W. Sherwood, John H. Alexander, Sunil V. Rao, J. Dawn Abbott, Jeffrey L. Carson, Andrew M. Goldsweig

## Abstract

**Background:** Although anemia is common in patients with myocardial infarction (MI), management remains controversial. We quantified the association of anemia with in-hospital outcomes and resource utilization in patients admitted with MI using a large national database.

**Methods:** All hospitalizations with a primary diagnosis code for acute MI in the National Inpatient Sample (NIS) between 2014 and 2018 were identified. Among these hospitalizations, patients with anemia were identified using a secondary diagnosis code. Data on demographic and clinical variables were collected. Outcomes of interest included in-hospital adverse events, length of stay (LOS), and total cost. Multivariable logistic regression and generalized linear models were used to evaluate the relationship between anemia and outcomes.

**Results:** Among 1,113,181 MI hospitalizations, 254,816 (22.8%) included concomitant anemia. Anemic patients were older and more likely to be women. After adjustment for demographics and comorbidities, anemia was associated with higher mortality (7.1 vs. 4.3%; odds ratio 1.09; 95% confidence interval [CI] 1.07-1.12, p<0.001). Anemia was also associated with a mean of 2.71 days longer LOS (average marginal effects [AME] 2.71; 95% CI 2.68-2.73, p<0.05), and higher total costs (mean cost $9,703 higher; AME $9703, 95% CI $9,577-$9,829, p<0.05). Anemic patients who received blood transfusions had higher mortality as compared with those who did not (8.2% vs. 7.0, p<0.001).

**Conclusion:** In MI patients, anemia was associated with higher in-hospital mortality, adverse events, total cost, and length of stay. Transfusion was associated with increased mortality, and its role in MI requires further research.

## Introduction

Coronary artery disease (CAD) is the leading cause of mortality in the United States, with approximately 805,000 people in the United States suffering a myocardial infarction (MI) every year.^1–3^ Previous studies have identified hypertension (52.3%), smoking (31.3%), dyslipidemia (28.0%), family history of CAD (28.0%), and diabetes (22.4%) as the most common risk factors in patients with MI.^4,5^ Anemia has also been recognized as an important comorbid condition in MI with smaller, older studies and studies of patients undergoing percutaneous coronary intervention (PCI) suggesting anemia as a risk factor for adverse outcomes.^6–8^

However, the relationship between anemia, adverse outcomes, and resource utilization in a broad population of patients hospitalized with MI remains incompletely characterized. Furthermore, management of these patients, in particular the effect of transfusion, remains controversial. The aim of the present study was to determine the association of anemia with in-hospital adverse outcomes, mortality, length of stay, and costs of care within the broad population of patients hospitalized with a MI using a large, nationally representative database.

## Methods

### Data Source

De-identified discharge-level data were obtained from the National Inpatient Sample (NIS) from 2014 through 2018. The NIS is the largest, all-payer inpatient database of the Agency for Healthcare Research and Quality’s (AHRQ’s) Healthcare Cost and Utilization Project (HCUP), containing an approximately 20% stratified sample of discharges from all hospitals in 27 US states.^9^ NIS data include demographics, socioeconomic status, comorbidities, length of stay, discharge status, total charges, and hospital characteristics. Diagnoses (up to 40) and procedures (up to 15) are encoded using International Classification of Diseases, Clinical Modification codes (ICD-9-CM, 2012 through third quarter 2015; ICD-10-CM from fourth quarter 2015 through 2018). We linked the NIS with cost-to-charge ratio files from the Healthcare Cost Report Information System to convert total charges to total costs.^10^ Costs reflect actual expenses incurred during production of hospital services such as wages, supplies and utility costs, whereas charges represent the amount a hospital billed for the hospitalization. We adjusted total costs for each year to 2018 US dollars ($) using the medical care consumer price index.^11^

### Study Population

The study population included all patients hospitalized with a primary diagnosis of MI, either non-ST-elevation myocardial infarction (NSTEMI) or ST-elevation myocardial infarction (STEMI), using ICD-CM codes presented in **Supplementary Table 1**.

This study was exempt from the requirements of the Institutional Review Board of the University of Nebraska Medical Center because the NIS contains no patient-identifiable information.

### Outcomes and Independent Variables

We studied two sets of outcomes: inpatient adverse events and mortality, and resource utilization in terms of length of stay (LOS) and total costs. A binary variable of any adverse events was constructed using seven adverse events including acute kidney injury (AKI), cardiac arrest, cardiogenic shock, hemorrhagic stroke, ischemic stroke, transient ischemic attack (TIA), and acute heart failure (HF) (ICD-CM codes in **Supplementary Table 1**).

The primary independent variable was a binary variable of anemia, which was identified by iron deficiency anemia, anemia of chronic disease, acute blood loss anemia, and other anemias. Corresponding ICD-CM codes are presented in **Supplementary Table 1**.

### Explanatory Variables

Explanatory variables included patient age (≤50, 51-60, 61-70, 71-80, 81-90), sex, Elixhauser comorbidity index, seven binary variables of health risk factors and health history (smoking, dyslipidemia, history of stroke/TIA, history of MI, history of CAD, atrial fibrillation, atrial flutter), and weekend admission. Age and Elixhauser comorbidity index were converted to categorical variables because their relationships with outcomes were highly non-linear; all other predictors were presented as categorical variables by the NIS. Using 3M All Patient Refined Diagnosis Related Groups, NIS, classifies severity of illness into minor (including cases with no comorbidity or complications), moderate, major, and extreme loss of function.^12^ Similarly, risk of mortality is categorized into minor, moderate, major, and extreme likelihood of dying.

Other sociodemographic variables included patient insurance status (Medicare, Medicaid, private, self-pay, no charge/other), neighborhood median household income quartile per ZIP code per year as a proxy of patient socioeconomic status, and county population density (≤249,999, 250,000-999,999, fringe counties of ≥1 million, central counties of ≥1 million population) as a proxy of urban/rural location.

Hospital characteristics assessed included ownership (for-profit private; not-for-profit private; government, non-federal), size (small, medium, large, per NIS criteria by region) and teaching status (urban teaching, urban non-teaching and rural).^13^

### Statistical Analysis

We examined differences in each variable according to anemia using Student’s t-tests and Pearson’s Chi-squared tests. To estimate the impact of anemia on inpatient LOS and total costs, we used a multivariable generalized linear model with a log link function and a gamma distribution (GLM-LG).^14,15^ Given that outcomes were skewed to the right and included outliers, GLM-LG made the distribution of outcomes approximately normal and estimated effects without bias. For inpatient complications and mortality, we used multivariable logistic models.

The estimation coefficients of GLM-LG were converted into average marginal effects (AME), which represent the differences in adjusted predicted outcome between a comparison group versus a reference group (i.e., with and without anemia). AME allows us to interpret estimates in terms of the outcome value (i.e., days for LOS, dollars for cost). AME is statistically significant (p<0.05) if the 95% confidence interval [CI] does not include zero. A sensitivity analysis was performed estimating total charges instead of total costs.

In the study cohort of patients who had anemia, we also examined the association of RBC transfusion with inpatient resource utilization and inpatient health outcomes, adjusting all other explanatory variables listed above. These patients undergoing RBC transfusion were identified based on ICD-9 and 10 procedure codes (**Supplementary Table 1)**.

All analyses were conducted with Stata MP v.16.1 and accounted for the discharge weighting in the HCUP NRD/NIS survey design in order to produce nationally representative estimates.

## Results

### Baseline Demographics

The study population included 1,113,181 patients with MI of whom 254,816 (22.8%) had comorbid anemia. Patient characteristics are presented in **Table 1**. Notably, the average age was 67.1 years, 38.1% were female, average Elixhauser comorbidity index was 2.2 (standard deviation [SD] 2.6), 42.5% had major or extreme loss of function due to severity of illness, and 44.3% had major or extreme likelihood of dying. Medicare, Medicaid, and private insurance covered 58.4%, 8.8%, and 24.9% of patients, respectively.

**Table 1.** Patient, Hospital, and Neighborhood Characteristics Among Total Study Population and Stratified by Anemia (No. (%))

|  | Total study | Anemia |  | p-value |
| --- | --- | --- | --- | --- |
|  | population<br>(n = 1,113,181) | No<br>(n = 858,365) | Yes<br>(n = 254,816) |  |
| Anemia | 254,816 (22.8) | 0 (0.0) | 254,816 (100.0) | < 0.001 |
| Age category |  |  |  | < 0.001 |
| ≤ 50 | 131,503 (11.8) | 114,331 (13.3) | 17,135 (6.7) | - |
| 51-60 | 230,965 (20.7) | 194,120 (22.6) | 36,800 (14.4) | - |
| 61-70 | 288,699 (25.9) | 223,922 (26.1) | 64,773 (25.4) | - |
| 71-80 | 244,663 (22.0) | 174,431 (20.3) | 70,273 (27.6) | - |
| 81-90 | 217,349 (19.5) | 151,559 (17.7) | 65,835 (25.8) | - |
| Women | 424,148 (38.1) | 312,068 (36.4) | 112,123 (44.0) | - |
| Elixhauser comorbidity index |  |  |  | < 0.001 |
| 0 | 386,273 (34.7) | 304,425 (35.5) | 81,829 (32.1) | - |
| 1-2 | 202,963 (18.2) | 189,277 (22.1) | 13,593 (5.3) | - |
| 3-4 | 248,931 (22.4) | 204,074 (23.8) | 44,823 (17.6) | - |
| 5-6 | 175,883 (15.8) | 114,358 (13.3) | 61,585 (24.2) | - |
| ≥ 7 | 99,131 (8.9) | 46,231 (5.4) | 52,985 (20.8) | - |
| Severity of illness |  |  |  | < 0.001 |
| Minor loss of function | 210,216 (18.9) | 205,025 (23.9) | 5,070 (2.0) | - |
| Moderate loss of function | 429,623 (38.6) | 364,849 (42.5) | 64,678 (25.4) | - |
| Major loss of function | 311,506 (28.0) | 195,948 (22.8) | 115,683 (45.4) | - |
| Extreme loss of function | 161,836 (14.5) | 92,543 (10.8) | 69,385 (27.2) | - |
| Risk of mortality |  |  |  | < 0.001 |
| Minor likelihood of dying | 292,249 (26.3) | 280,978 (32.7) | 11,114 (4.4) | - |
| Moderate likelihood of dying | 328,444 (29.5) | 269,650 (31.4) | 58,748 (23.1) | - |
| Major likelihood of dying | 321,404 (28.9) | 207,931 (24.2) | 113,585 (44.6) | - |
| Extreme likelihood of dying | 171,083 (15.4) | 99,805 (11.6) | 71,369 (28.0) | - |
| Smoking/tobacco use | 353,630 (31.8) | 291,794 (34.0) | 61,781 (24.2) | < 0.001 |
| Dyslipidemia | 736,108 (66.1) | 567,139 (66.1) | 168,971 (66.3) | 0.006 |
| History of stroke/TIA | 34,143 (3.1) | 24,275 (2.8) | 9,873 (3.9) | < 0.001 |
| History of MI | 166,621 (15.0) | 124,498 (14.5) | 42,134 (16.5) | < 0.001 |
| History of CAD | 919,278 (82.6) | 703,553 (82.0) | 215,740 (84.7) | < 0.001 |
| Atrial fibrillation | 211,310 (19.0) | 140,123 (16.3) | 71,252 (28.0) | < 0.001 |
| Atrial flutter | 25,639 (2.3) | 16,946 (2.0) | 8,701 (3.4) | < 0.001 |
| Weekend admission | 297,770 (26.7) | 231,572 (27.0) | 6,6193 (26.0) | < 0.001 |
| Insurance status |  |  |  | < 0.001 |
| Medicare | 649,722 (58.4) | 467,059 (54.4) | 182,760 (71.7) | - |
| Medicaid | 98,373 (8.8) | 79,008 (9.2) | 19,356 (7.6) | - |
| Private | 277,453 (24.9) | 237,696 (27.7) | 39,690 (15.6) | - |
| Self-pay | 50,326 (4.5) | 43,842 (5.1) | 6,470 (2.5) | - |
| No charge/other | 37,307 (3.4) | 30,761 (3.6) | 6,540 (2.6) | - |
| Neighborhood income |  |  |  | < 0.001 |
| Bottom quartile | 336,997 (30.3) | 256,661 (29.9) | 80,345 (31.5) | - |
| Second quartile | 311,934 (28.0) | 241,649 (28.2) | 70,281 (27.6) | - |
| Third quartile | 264,805 (23.8) | 205,268 (23.9) | 59,534 (23.4) | - |
| Top quartile | 199,446 (17.9) | 154,787 (18.0) | 44,655 (17.5) | - |
| Neighborhood urban-rural |  |  |  | < 0.001 |
| Counties ≤ 249,999 | 349,577 (31.4) | 275,566 (32.1) | 73,993 (29.0) | - |
| Counties 250,000 - 999,999 | 236,767 (21.3) | 186,083 (21.7) | 50,674 (19.9) | - |
| Fringe counties, ≥ 1 million | 269,623 (24.2) | 206,484 (24.1) | 63,142 (24.8) | - |
| Central counties, ≥ 1 million | 257,214 (23.1) | 19,0231 (22.2) | 67,006 (26.3) | - |
| Hospital ownership |  |  |  | < 0.001 |
| Private, investor-owned | 176,336 (15.8) | 137,145 (16.0) | 39,188 (15.4) | - |
| Private, not-for-profit | 831,375 (74.7) | 638,715 (74.4) | 192,667 (75.6) | - |
| Government, non-federal | 105,469 (9.5) | 82,505 (9.6) | 22,961 (9.0) | - |
| Hospital size |  |  |  | < 0.001 |
| Small | 163,631 (14.7) | 128,794 (15.0) | 34,829 (13.7) | - |
| Medium | 322,719 (29.0) | 251,344 (29.3) | 71,368 (28.0) | - |
| Large | 626,831 (56.3) | 478,227 (55.7) | 148,619 (58.3) | - |
| Hospital teaching status |  |  |  | < 0.001 |
| Urban, non-teaching | 289,784 (26.0) | 228,421 (26.6) | 61,349 (24.1) | - |
| Non-metropolitan | 83,953 (7.5) | 67,923 (7.9) | 16,020 (6.3) | - |
| Urban, teaching | 739,444 (66.4) | 562,021 (65.5) | 177,446 (69.6) | - |
| Year |  |  |  | 0.004 |
| 2014 | 200,935 (18.1) | 156,080 (18.2) | 44,851 (17.6) | - |
| 2015 | 212,607 (19.1) | 163,857 (19.1) | 48,751 (19.1) | - |
| 2016 | 219,346 (19.7) | 168,622 (19.6) | 50,725 (19.9) | - |
| 2017 | 240,706 (21.6) | 185,346 (21.6) | 55,360 (21.7) | - |
| 2018 | 239,587 (21.5) | 184,459 (21.5) | 55,128 (21.6) | - |

Note: percentages were adjusted for discharge weight to generate nationally representative individual level estimates.

### Adverse Events, Mortality, LOS, and Costs

Among the total study population, composite rate of adverse events was 34.5 %, and in-hospital mortality was 5.0%. The adverse event rate (55.1% vs. 28.4%) and mortality rate (7.1% vs 4.3%) were higher among patients with anemia than among patients without anemia (p<0.001 for all, **Table 2**). AKI and HF constituted majority of adverse events experienced by the overall study population. Furthermore, the incidence of both AKI (35.1 vs 14.3, p<0.001) and HF (31.4 vs 15.7, p<0.001) were higher in anemic patients. The average inpatient LOS was 4.63 ± 5.63 days, and the average total cost was $23,046 ± 24,544. Average LOS (7.93 vs. 3.66 days) and cost ($33,541 vs. $19,939) were higher among patients with anemia than without anemia.

**Table 2.** Inpatient Resource Utilization and Inpatient Health Outcomes among Total Study Population and Stratified by Anemia

|  | Total study | Anemia |  | p-value |
| --- | --- | --- | --- | --- |
|  | population<br>(n = 1,113,181) | No<br>(n = 858,365) | Yes<br>(n = 254,816) |  |
| Length of stay (days), mean (SD) | 4.63 (5.63) | 3.66 (4.27) | 7.93 (7.95) | < 0.001 |
| Total cost (\$), mean (SD) | 23,046 (24,544) | 19,939 (18,381) | 33,541 (36,789) | < 0.001 |
| Total charge (\$), mean (SD) | 96,537 (118,241) | 82,724 (90,720) | 143,199 (174,890) | < 0.001 |
| Adverse Events, n (%) |  |  |  |  |
| Any | 383,575 (34.5) | 243,350 (28.4) | 140,373 (55.1) | < 0.001 |
| Acute kidney injury | 211,755 (19.0) | 122,490 (14.3) | 89,380 (35.1) | < 0.001 |
| Cardiac arrest | 36,593 (3.3) | 25,514 (3.0) | 11,087 (4.4) | < 0.001 |
| Cardiogenic shock | 49,743 (4.5) | 28,857 (3.4) | 20,913 (8.2) | < 0.001 |
| Hemorrhagic stroke | 1,067 (0.1) | 692 (0.1) | 375 (0.1) | < 0.001 |
| Ischemic stroke | 13,777 (1.2) | 8,741 (1.0) | 5,041 (2.0) | < 0.001 |
| Transient Ischemic Attack | 4,024 (0.4) | 2,918 (0.3) | 1,106 (0.4) | < 0.001 |
| Acute heart failure | 214,379 (19.3) | 134,353 (15.7) | 80,114 (31.4) | < 0.001 |
| Mortality, n (%) | 55,320 (5.0) | 37,192 (4.3) | 18,143 (7.1) | < 0.001 |

Note: days, costs, charges, standard deviations (SD), and percentages were adjusted for discharge weight to generate nationally representative individual level estimates.

Notably, average LOS continuously decreased over time (4.88 days in 2014 to 4.40 days in 2018), and total costs remained relatively stable over the 5-year study period ($23,972 in 2014 to $22,316 in 2018). However, total charges increased ($92,364 in 2014 to $100,373 in 2018), even when adjusted for the medical care consumer price index (**Supplementary Table 2**).

### Multivariable Analyses

In multivariable analyses for adverse outcomes (**Table 3**), anemia was associated with an increased risk for adverse outcomes (odds ratio [OR] 1.89; 95% CI 1.87 to 1.91) and in-hospital mortality (OR 1.09; 95% CI 1.07 to 1.12) compared with patients without anemia after adjustment for measured confounders. These associations between anemia and adverse outcomes, and anemia and mortality were significant among all age groups (**Supplementary Table 3**).

**Table 3.** Multi-variable Analysis of Anemia and Inpatient Adverse Outcomes, Mortality and Resource Utilization

|  |  |
| --- | --- |
| In-Hospital Adverse Events, OR (95% CI) | 1.09 (1.07 – 1.12) |
| In-Hospital Mortality, OR (95% CI) | 1.89 (1.87 – 1.91) |
| Length of Stay, AME (95% CI) | 2.71 days (2.68 – 2.73) |
| Total Costs, AME (95% CI) | \$ 9,703 (9,577 – 9,829) |
| Total Charges, AME (95% CI) | \$ 41,967 (41,388 – 42,547) |

Notes:
(i) Odds ratio (OR) and 95% confidence interval (CI) were adjusted for discharge weight to generate nationally representative individual level estimates.
(ii) Average marginal effect (AME) and 95% confidence interval (CI) were adjusted for discharge weight to generate nationally representative individual level estimates.
(iii) AME is statistically significant ( $p < 0.05$ ) if 95% CI does not include zero.

In multivariable analyses for resource utilization, compared with no anemia, anemia was associated with a longer average LOS by 2.71 days (AME 2.71; 95% CI: 2.68 to 2.73), and higher total costs by $9,703 (AME 9,703, 95% CI: 9,577 to 9,829) (**Table 3**).

High Elixhauser comorbidity index, large hospital size, and urban teaching hospital were consistently associated with higher rates of adverse outcomes and resource utilization (**Supplementary Tables 3 and 4**).

### Sensitivity Analysis

In the sensitivity analysis examining total charges, the average total charge was $96,537 ± 118,241 and was significantly higher among patients with anemia ($143,199 vs. $82,724; p < 0.001) (**Table 2**). Compared with patients having no anemia, total charges were higher by $41,967 (AME 41,967; 95% CI 41,388 to 42,547) among patients with anemia in multivariable analysis adjusting for all explanatory variables (**Table 3**).

### Blood Transfusions in Patients with Anemia

Among patients with anemia, 46,645 (18.3%) received a blood transfusion. These patients had higher rates of adverse events (62.4% vs. 53.6%) and mortality (8.4% vs. 6.9%, p<0.001) than patients with anemia who did not receive a blood transfusion (**Table 4**). Furthermore, blood transfusion in patients with anemia was associated with increased length of stay (mean [SD]), 10.69 (8.91) vs. 7.37 (7.62) days and total costs (mean [SD]), $44,100 (38,886) vs. $31,400 (35,973).

**Table 4.** Effect of RBC Transfusion on Inpatient Adverse Events, Mortality and Resource Utilization (n = 254,816)

|  | RBC transfusion |  | p-value |
| --- | --- | --- | --- |
|  | No<br>(n = 208,171) | Yes<br>(n = 46,645) |  |
| Length of stay (days), mean (SD) | 7.37 (7.62) | 10.69 (8.91) | < 0.001 |
| Total cost (\$), mean (SD) | 31,400 (35,973) | 44,100 (38,886) | < 0.001 |
| Total charge (\$), mean (SD) | 134,212 (170,286) | 187,519 (189,898) | < 0.001 |
| Adverse Events, n (%) |  |  |  |
| Any | 111,586 (53.6) | 29,112 (62.4) | < 0.001 |
| Acute kidney injury | 69,939 (33.6) | 19,765 (42.4) | < 0.001 |
| Cardiac arrest | 8,688 (4.2) | 2,437 (5.2) | < 0.001 |
| Cardiogenic shock | 16,472 (7.9) | 4,506 (9.7) | < 0.001 |
| Hemorrhagic stroke | 300 (0.1) | 76 (0.2) | < 0.001 |
| Ischemic stroke | 3,901 (1.9) | 1,163 (2.5) | < 0.001 |
| TIA | 882 (0.4) | 227 (0.5) | < 0.001 |
| Acute heart failure | 63,960 (30.7) | 16,310 (35.0) | < 0.001 |
| Mortality, n (%) | 14,261 (6.9) | 3,941 (8.4) | < 0.001 |

Note: days, costs, charges, standard deviations (SD), and percentages were adjusted for discharge weight to generate nationally representative individual level estimates.

## Discussion

In this analysis of over 1 million patients hospitalized with MI, we found that anemia was associated with more in-hospital adverse events, higher mortality, longer length of stay, and higher total costs. Furthermore, blood transfusion among anemic MI patients was associated with worse outcomes and increased resource utilization.

### Adverse Events

Anemia among MI patients was associated with both a composite of seven adverse events (AKI, cardiac arrest, cardiogenic shock, hemorrhagic stroke, ischemic stroke, TIA, and acute HF) as well as with each of those events individually. However, two components of these adverse events primarily drove the composite, affecting more than 10% of patients both with and without anemia: AKI and acute HF.

### Mechanisms of Adverse Events

Both pathophysiologic and clinical mechanisms may explain the higher rates of adverse outcomes among MI patients with anemia. Pathophysiologically, oxygen delivery depends upon hemoglobin (Hb) and cardiac output. In the setting of MI, anemia worsens ischemia by decreasing oxygen delivery to the affected myocardium and increasing myocardial oxygen demand due to greater cardiac output required to maintain adequate systemic perfusion.^16^ Indeed, when tissue oxygen needs are not met adequately, reflex increases in heart rate and stroke volume to increase cardiac output result in increased cardiac work and thereby worsen myocardial oxygen supply-demand mismatch.^17^ Anemia may also be a marker of comorbidities that worsen MI outcomes such as diabetes, chronic kidney disease, and active bleeding.^18,19^

Clinically, patients with anemia may be less likely to receive antiplatelet and anticoagulant therapies due to bleeding concerns, which could impact cardiovascular events and mortality. For, example, in the CADILLAC (Controlled Abciximab and Device Investigation to Lower Late Angioplasty Complications) trial, 18% of patients with anemia at the time of ACS were no longer taking aspirin at 1 year follow-up.^6^ Also, patients with anemia are less likely to undergo PCI, as seen in the ACUITY (Acute Catheterization and Urgent Intervention Triage Strategy) trial. These disparities in care may contribute to worse cardiovascular outcomes. ^20^

The incidence of AKI among MI patients was more than double in the presence of anemia. Anemia plays an important role in the worsening of renal function in acute coronary syndromes (ACS).^21,22^ Anemia reduces the oxygen carrying capacity of blood, which may lead to renal damage by aggravating ischemia and hypoxia of the renal medulla.^23^ Anemia is also associated with an increased incidence of contrast-associated AKI in patients undergoing coronary angiography.^24^ Anemia has been identified as an independent predictor for development of AKI in patients undergoing PCI by multiple risk prediction models.^25 26^ Additionally, in patients with underlying HF, anemia can further exacerbate HF, leading to renal injury, the so call cardio-renal-anemia syndrome.^27^

Furthermore, AKI has previously been associated with higher mortality risk in patients with MI.^28–30^ Kuno et al. reported that patients undergoing PCI with a > 3 g/dL decrease in peri-procedural Hb level or a 20% relative decrease in Hb had significantly increased rates of AKI associated with increased in-hospital mortality.^31^

Similar to AKI, the incidence of acute HF was doubled in the presence of anemia. Anemia reduces oxygen delivery to myocardium downstream of coronary stenoses, leading to impaired myocardial healing and an exaggerated inflammatory response.^8,32,33^ The combination of these processes exacerbates HF in this population. Our findings concur with those of Kruk et al., who found significantly worse HF in patients with MI and anemia compared to those without anemia.^27^ Among patients with ACS, HF progresses as Hgb drops below 14 g/dL.^7^ A cohort study by Archbold et al showed that anemia is a powerful independent determinant of left ventricular systolic dysfunction in patients with ACS.^34^ The rate of HF was inversely related to Hb concentration, with increasing frequency of HF as Hb levels decreased. Additionally, anemia has also been shown to be an independent risk factor for adverse cardiovascular outcomes in patients with HF.^35,36^

### Mortality

We found that in-hospital mortality was significantly higher in MI patients with anemia than without anemia (7.2% vs. 4.6%, p<0.001). Many prior studies have reported conflicting findings with regard to the relationship between anemia and mortality in patients with ACS.^19,37– 40^ Mamas et al, retrospectively analyzing the MINAP (Myocardial Ischemia National Audit Project) registry between January 2006 and December 2010, found an independent association between anemia and a ∼50% higher risk of short- and long-term mortality in both men and women.^41^ Similarly, in a meta-analysis by Lawler et al that included 27 studies (233,144 patients), anemia was associated with an increased risk of crude all-cause mortality (relative risk 2.08, 95% CI 1.70 to 2.55) at longest available follow-up.^42^

Conversely, other studies have reported that, with adjustment for demographics and comorbidities, anemia is no longer an independent predictor of mortality.^7,43^ The HORIZONS-AMI (Harmonizing Outcomes with Revascularization and Stents in Acute Myocardial Infarction) trial reported different relationships between anemia and cardiovascular outcomes according to sex, with anemia independently associated with higher rates of all-cause and cardiac mortality at 30 days and 1 year in men but not in women.^40^ The risk for adverse cardiovascular events may also be higher with ACS and elevated baseline Hb (>16 to 17 g/dL), perhaps as a marker of chronic hypoxia.^44^

### Length of Stay and Total Cost

Given the worse in-hospital clinical outcomes associated with anemia in patients with MI, it is not surprising that LOS and total hospital costs were significantly higher in patients with anemia. Further research is warranted to develop and validate innovative, patient-centered strategies to reduce adverse outcomes and thus LOS and cost associated with anemia in patients admitted with MI.

### Transfusion

Our analysis also showed that RBC transfusion in MI patients with anemia was associated with increased adverse outcomes and mortality. The observational methodology precludes determining whether this relationship is causative or merely represents increased use of transfusion in sicker patients. Indeed, the role of transfusion in MI remains a contentious topic of uncertainty and on-going research. In a pooled analysis of 24,112 patients in 3 large ACS trials, transfusion was associated with increased mortality (adjusted hazard ratio 3.94; 95% CI 3.26-4.75).^45^ Conversely, a retrospective study of 78,974 Medicare beneficiaries with MI associated transfusion with decreased risk of mortality.^46^ In the middle, the prospective randomized REALITY (Effect of a Restrictive vs. Liberal Blood Transfusion Strategy on Major Cardiovascular Events Among Patients with Acute Myocardial Infarction and Anemia) trial found no 30-day mortality difference between patients with MI and anemia transfused to a Hb target of 8 or 10 g/dL.^47^ The on-going randomized 3500-patient MINT (Myocardial Ischemia and Transfusion), trial (NCT02981407) is designed to provide a more definitive answer regarding the role of transfusion in a broad population of patients with MI and anemia as well as several pre-specified subgroups.^48^

### Strengths and Limitations

This analysis utilized the NIS database, providing the largest study to date on the impact of anemia in MI. This sample size provided adequate power to detect differences in outcomes not evident in smaller studies. Hard endpoints such as adverse events, mortality, LOS, and costs are likely to be coded correctly. NIS data are geographically and ethnically diverse and nationally representative of real-world practice and outcomes. The data are subject to rigorous quality control, and NIS is well-validated for such outcomes.

However, the present study has several important limitations. First, in studies using administrative claims codes, any incorrect coding may lead to inaccurate and potentially missing data. Second, the retrospective nature introduces inherent selection bias. Third, given the administrative nature of database, detailed clinical characteristics such as patients’ baseline Hb were not available. Fourth, for the same reason, utilization of medical therapy, in particular antithrombotic medications, could not be assessed. Fifth, outcomes were confined to in-hospital events in this inpatient database, and long-term clinical outcomes remain unknown. Finally, data on the prevalence of baseline comorbid conditions such as diabetes, hypertension, chronic HF, and chronic kidney disease were not included in the NIS database starting in the fourth quarter of 2015 through 2018. This limitation, reflecting the administrative nature of the database, occurred with transition from ICD-9 codes to ICD-10 codes. Thus, these variables could not be included in multivariable logistic models used for our analysis.

## Conclusion

In patients admitted with MI, anemia was associated with more adverse events, higher mortality, longer length of stay, and higher total cost. Transfusion was associated with increased adverse events and mortality, and its role in patients with MI and anemia requires further research.

## Data Availability

The authors confirm that the data supporting the findings of this study are available within the article [and/or] its supplementary materials.

---

**Supplementary Table 1**. International Classification Disease (ICD) Codes for Primary Variables.

**Supplementary Table 2**. Inpatient Resource Utilization and Inpatient Health Outcomes over Years.

**Supplementary Table 3**. Multi-variable Analysis of Association of Anemia with Inpatient Adverse Events and Mortality stratified by different sub-groups.

**Supplementary Table 4**. Multi-variable Analysis of Association of Anemia with Inpatient Length of Stay, Total Cost, and Total Charges stratified by different sub-groups.

